# Genome-wide Association Study of Postpartum Depression Identifies a Novel Susceptibility Locus at 18q12.1

**DOI:** 10.1101/2023.04.24.23289058

**Authors:** Aldo Cordova-Palomera, Dorothée Diogo, Sándor Szalma

## Abstract

Postpartum depression (PPD) is among the most frequent and incapacitating conditions following childbirth, with significant consequences for mothers, newborns and families. Genetic factors have been proposed to influence disease risk and symptom heterogeneity, and can potentially inform drug target discovery and treatment strategies.

Here, we conducted a genetic association study to further our understanding of the genetic architecture of PPD. We identified PPD cases and controls in the UK Biobank using multiple sources of medical history and self-reported information. We performed genome-wide association studies of common and rare variants in in a harmonized set of up to 11,782 PPD cases and 167,480 controls among European-ancestry females.

Genetic association results displayed a significantly associated locus at chromosome 18q12.1 led by the common rs10502503 marker (minor allele frequency: 29.9%, effect allele: C, odds ratio: 0.92, *p*=6.4×10^−9^), with *in silico* functional mapping suggesting Cadherin 2 (*CDH2*) as a candidate causal gene. This signal, if confirmed in independent replication cohorts with PPD diagnosis confirmed through psychometry, may contribute novel insights into the genetic basis of PPD.

The results illustrate the use of minimal phenotyping in large-scale general population cohorts to investigate the genetic etiology and heterogeneity of PPD, and to generate therapeutic hypotheses.

## Introduction

With an overall prevalence rate around 17% in women worldwide [1] and ostensibly higher estimates during the COVID-19 pandemic [2], postpartum depression (PPD) is a debilitating mental condition that can significantly impact health and wellbeing of mothers, children, infants and spouses. Risk factors frequently reported in the epidemiological literature include maternal age, gestational diabetes, depression history and psychosocial adversities [3, 4], and treatment strategies comprise psychosocial, psychotherapeutic, pharmacological and even somatic interventions [5]. Central to this diversity of premorbid conditions and treatment strategies, PPD encompasses a wide range of clinical outcomes, and heterogeneous subtypes can be distinguished on the basis of e.g., severity, onset timing and comorbidities, with potential implications for phenotyping and genetic studies [6, 7]. In this regard, research on the genetic architecture of PPD have the potential to inform on disease subtypes, susceptibility factors beyond well-established epidemiological risk, and candidate biological pathways that can translate into safe drug treatment options to assist conventional non-pharmacological strategies.

Early quantitative genetics data from a twin study by Treloar and colleagues [8] indicated that PPD has a substantial genetic component that could be even larger than for other depressive disorders: a heritability estimate of 38% for PPD vs. 25% for the non-perinatal phenotype, and low genetic correlation between those traits (r(g) = 0.17). Consistently, a more recent twin study by Viktorin and colleagues [9] reported higher heritability for PPD (44-54%) than for the non-perinatal phenotype (32%), and one-third of PPD’s genetic architecture not shared with non-perinatal depression.

Expanding on this genetic component of PPD first identified in twin cohorts, efforts to investigate the genetics of PPD using molecular genetics tools are underway: analysis of common genetic variants indicates a clear narrow-sense (SNP-based) heritability component around 22%, and low genetic overlap with other affective disorders [10]. Of note, recent work published by Kiewa and colleagues [11] encompassed a genome-wide association study (GWAS) of 3,804 well-characterized PPD cases and 6,134 controls from the Australian Genetics of Depression Study 2018 identified through the Edinburgh Postnatal Depression Scale [12, 13], which did not identify genomic loci meeting statistical significance criteria for association. To the best of authors’ knowledge, the latter is the largest PPD GWAS available to date in the literature. In this study, we aimed at getting further insights into the genetic architecture of PPD, by performing a GWAS of common variants (minor allele frequency ≥ 0.01) and a rare-variant association study (RVAS) (minor allele frequency ≤ 0.001) in a harmonized set of up to 11,782 PPD cases and 167,480 controls among European-ancestry females phenotyped through a pipeline combining medical records, hospital encounter data and self-reported questionnaires from the UK Biobank.

## Methods

### Participants and ethics statement

Data for this PPD analysis was retrieved from the UK Biobank (application 26041), a longitudinal cohort study of >500,000 participants aged 40-69 years enrolled in study centers across England, Wales and Scotland between 2006 and 2019. In addition to extensive baseline data about medical histories, physical measurements and health behavior of participants, biosamples were collected from the participants (e.g., saliva, urine and blood). Ethical approval was granted for the UK Biobank by the North West Multi-Centre Research Ethics Committee and the National Health Service (NHS) National Research Ethics Service (ref: 11/NW/0382), all participants provided written informed consent, and all experiments were conducted in accordance to relevant regulations and guidelines. Additional information, including details on study protocol, are available online at https://biobank.ctsu.ox.ac.uk/.

### Case-control definitions

Definitions of PPD cases and controls, as well as exclusion criteria were obtained by leveraging information from demographics, medical histories, hospital encounters, general practitioner records and self-reported questionnaires. Longitudinal data was aligned to the International Classification of Diseases, tenth version (ICD-10) ontology [14] following dedicated data curation, cleanup and quality control practices outlined elsewhere (https://ranchobiosciences.com/services/). In the analyses described below, when referring to medical data sources (hospital encounters, medical histories and general practitioner data), phenotyping was done using ICD-10 codes: cases were identified using the F53* prefix (mental and behavioural disorders associated with the puerperium, not elsewhere classified); controls were selected using O00-O9A codes (pregnancy, childbirth and the puerperium) and certain Z32* to Z39* prefixes (Z32 - encounter for pregnancy test and childbirth and childcare instruction; Z33 - pregnant state; Z34 - encounter for supervision of normal pregnancy; Z36 - encounter for antenatal screening of mother; Z37 - outcome of delivery; Z38 - liveborn infants according to place of birth and type of delivery; Z39 - encounter for maternal postpartum care and examination). Identification of cases and controls consisted of the following stages:

1. excluding all male participants;
2. retrieving case-control labels from medical data sources using UK Biobank data-fields 40001, 40002, 40006, 40013, 41270, 41271, 20001, 20002, 20544 and GP diagnoses (n_cases=8,958, n_controls=78,333);
3. expanding the pool of controls with data-field 41261, “records in HES inpatient maternity dataset” (cumulative n_cases=8,958, cumulative n_controls=79,445);
4. expanding the pool of cases with data-field 20445, “depression possibly related to childbirth, self-reported” (cumulative n_cases=14,810, cumulative n_controls=79,445); and
5. expanding the pool of controls by including women with at least one live birth (self-reported, as retrieved from data-field 2734) (cumulative n_cases=14,810, cumulative n_controls=217,199).

### Genetic association analysis of common variants (GWAS)

Cases and controls from the last stage above (14,810 cases and 217,199 controls) were further restricted to genetically unrelated women of European ancestry to conduct a GWAS of common variants in a sample of up to 11,782 cases and 167,480 controls, as described next. Genotypic data collected through either the Affymetrix Axiom Array or the custom UK Biobank Axiom array and imputed to the Haplotype Reference Consortium [15] and the joint UK10K/1000 Genomes panels was accessed through the UK Biobank data showcase.

Association statistics were calculated for imputed genotypes with a minimum minor allele frequency threshold of 0.01 and an empirical-theoretical variance ratio (MaCH’s *r*^2^) between 0.3 and 1.0. Analyses were performed on the largest available subset of unrelated PPD cases and controls of European ancestry as indexed by a kinship coefficient of at least 0.0844 (third degree relationship). Tests were conducted using standard logistic regression as implemented in PLINK (v2.00a2.3LM 64-bit Intel (24 Jan 2020)) [16], including age and the first two genetic principal components as covariates to control for population stratification and cryptic relatedness. Results from this stage are presented on hg19 coordinates.

### Statistical fine-mapping of GWAS results

When relevant, SNPs in a 1-megabase window (+-500kb) around the lead variant were submitted to Bayesian genetic fine-mapping, a procedure aimed at identifying causal variants within a locus responsible for an association. This analysis outputs credible sets of plausible causal variants that are then interpreted as including the causal variant at some probability [17]. Here, Bayesian fine-mapping was implemented using the *corrcoverage* package (https://github.com/annahutch/corrcoverage; https://cran.r-project.org/web/packages/corrcoverage/index.html).

### Functional annotation of GWAS results and Bayesian Tests for Colocalization

Annotation of genetic associations, gene prioritization and functional mapping were performed using FUMA, a platform to aid interpretation of GWAS results by automatic integration of diverse biological datasets [18]. In short, FUMA leverages positional, expression quantitative trait loci (eQTL) and chromatin interaction data to retrieve gene-based, pathway and tissue enrichment information. GWAS summary statistics are submitted to FUMA to compute gene-based *p*-values and gene set *p*-values using MAGMA [19], which performs regression models to improve statistical power optimizing computational performance. MAGMA maps markers to genes positionally, by assigning each marker to a gene based on whether it is inside the coordinates defined by transcription start and stop sites of that gene. All significant markers in linkage disequilibrium are then mapped to a catalog of eQTLs. With default parameters, only *cis*-eQTLs marker-gene pairs (mapping SNPs to genes up to 1 Mb apart) with significant false discovery rate *p* (under 0.05) were prioritized. FUMA also enables chromatin interactions mapping, where SNPs are mapped to genes when there is a significant chromatin interaction between the disease-associated regions and nearby or distant genes, using chromatin interaction data in various tissues and cell types. Those markers are then mapped to genes whose promoters overlap another end of interactions.

Expanding on the results retrieved from the FUMA platform, Bayesian colocalization tests were implemented to assess whether pairs of association signals were consistent with a shared causal variant [20]. In the current setting, summary statistics retrieved from a given PPD locus were contrasted against eQTL data for the same region extracted from FUMA. The *coloc* package in R (https://cran.r-project.org/web/packages/coloc/index.html) was used to run fully Bayesian colocalization analysis with Bayes factors, with PPD as trait 1 and default settings: prior for a marker associated with trait 1 equal to 10^−4^, prior for a marker associated with trait 2 equal to 10^−4^ and prior for a marker associated with both traits equal to 10^−5^.

### Genetic association analysis of rare variants (RVAS)

Exome sequencing data were generated by the UK Biobank Exome Sequencing Consortium (UKB-ESC) as described previously [21]. In brief, DNA samples were transferred from the UK Biobank to the Regeneron Genetics Center and stored in an automated sample biobank to undergo sample preparation and sequencing by means of a fully-automated, high-throughput approach developed at the Regeneron Genetics Center. Next, sample read mapping was conducted, followed by a stage of variant calling, aggregation and quality control [22], and an additional phase of low-quality variant identification using machine learning. Extended details on sample management, quality control and pre-processing of exome sequencing data can be found elsewhere [23].

Association tests for rare variants were conducted on the largest available set of unrelated European cases and controls using logistic regression tests, in PLINK (v2.00a2.3LM 64-bit Intel (24 Jan 2020)) [16], with the “firth-fallback” modifier to implement Firth regression when the standard test fails to converge, and controlling for age and the first two genetic principal components. Variants were filtered using a maximum minor allele frequency threshold of 0.001 in the entire case/control cohort and the analysis was restricted to markers annotated as high-confidence protein-truncating variants by the Variant Effect Predictor (VEP) [24] or the Loss-Of-Function Transcript Effect Estimator (LOFTEE) [25], or with high probability of pathogenicity as indexed by a score above 0.5 by the Helix Pathogenicity Prediction Platform [26]. In addition, gene collapse genotypes were computed by combining rare variants (minor allele frequency ≤ 0.001 with VEP, LOFTEE or Helix annotations); they were submitted to association testing after an additional filtering stage consisting to minor allele count in cases ≥ 20, as motivated above.

## Results

Table 1 summarizes age distributions of PPD cases and controls, as well as relevant psychosocial and clinical variables related to known PPD risk: socioeconomic status, employment type, educational level, other depression diagnoses, vitamin D and obesity (as reflected by body mass index). Statistically significant differences between PPD cases and controls were observed for several variables, indicative of an association between PPD risk and younger participant age, younger age at delivery, higher number of self-reported depression episodes lasting two weeks or longer, higher body mass index, lower overall vitamin D levels. Socioeconomic status variables displayed a pattern indicative of higher education, employment and income score in English PPD women, whereas the opposite trend was observed in Welsh and Scottish participants. A moderate correlation (*r*=0.52) was observed between participants’ age, and reported age at delivery (Supplementary Figure S1).

**Table 1.**
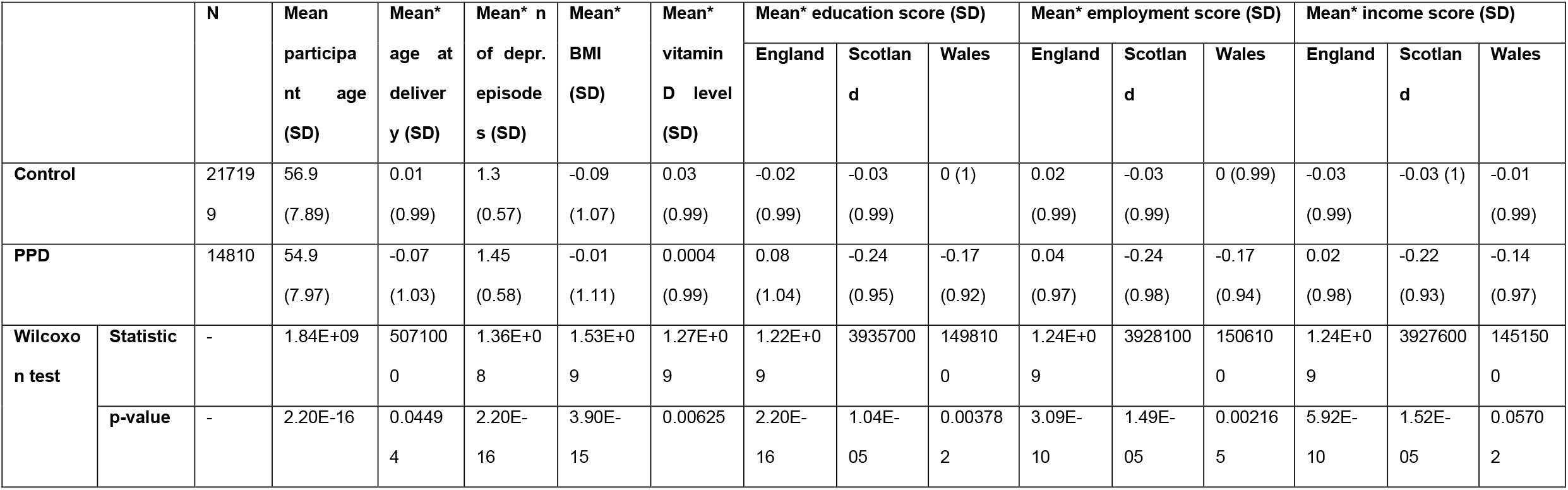
Demographic and risk factors for PPD, stratified by cases and controls. Notes: *, standardized variables (zero mean and unit standard deviation in the entire UKB).

PPD case and control labels were submitted to association tests for over 8.5 million markers using standard logistic regression. Summary statistics did not indicate substantial inflation (genomic inflation factor [λ], 1.09; Supplementary Figure S2) and revealed one genome-wide significant association (Figure 1) in a region near Cadherin 2 (*CDH2*) (locus coordinates: chr18:25322322-25559158; lead variant: rs10502503 [chr18:25413104], A1: C, AX: T, MAF in cases: 28.2%, MAF in controls: 29.98%; odds ratio: 0.92, β=-0.087; SE=0.015; *p*=6.4×10^−9^). As an additional check to rule out potential artifacts of genotype imputation, association tests were repeated for this locus using directly genotyped data; both outcomes were consistent (Supplementary Figure S3). The 90% credible set obtained through Bayesian fine mapping had a claimed coverage of 90.4% and included 11 variants (sorted decreasingly by posterior probability of causality: rs10502503, rs11662671, rs67108301, rs2002401, rs8092192, rs9959491, rs11876427, rs16944182, rs8093989, rs7243670, rs7235990; Supplementary Figure S4 and Supplementary Table S1). Submission of this 11-SNP set to the Ensembl Variant Effect Predictor [24] indicated that rs16944182 might have consequences on downstream gene regulation, upstream gene regulation and regulatory features of an open chromatin region; the remaining SNPs were annotated as intergenic, intronic or non-coding transcript variants. In general, linkage disequilibrium between the lead variant (rs10502503) and other markers in the region was not high (*R*^2^ < 0.8) (Figure 2); for the specific case of rs10502503 and rs16944182, pairwise linkage disequilibrium metrics across multiple European populations were estimated as D’ of 0.93 and *R*^2^ equal to 0.57 [27]. Posterior probabilities of causality from marginal Z-scores were 19.8% for the lead marker (rs10502503) and 2% for rs16944182.

**Figure 1.**
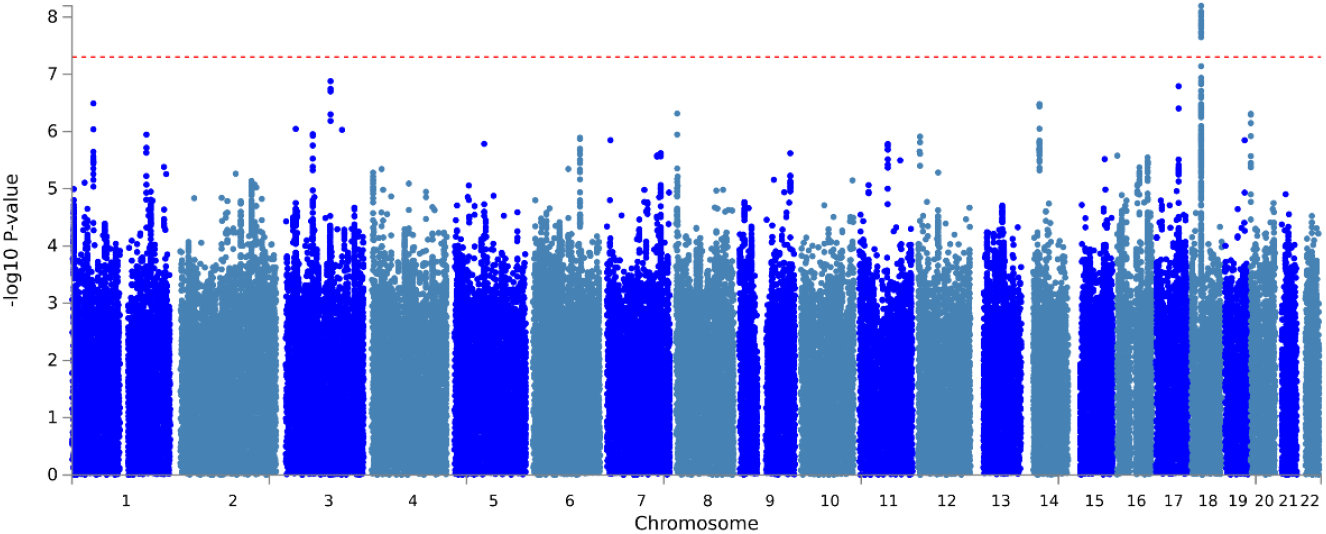
Manhattan plot for the GWAS of PPD.

**Figure 2.**
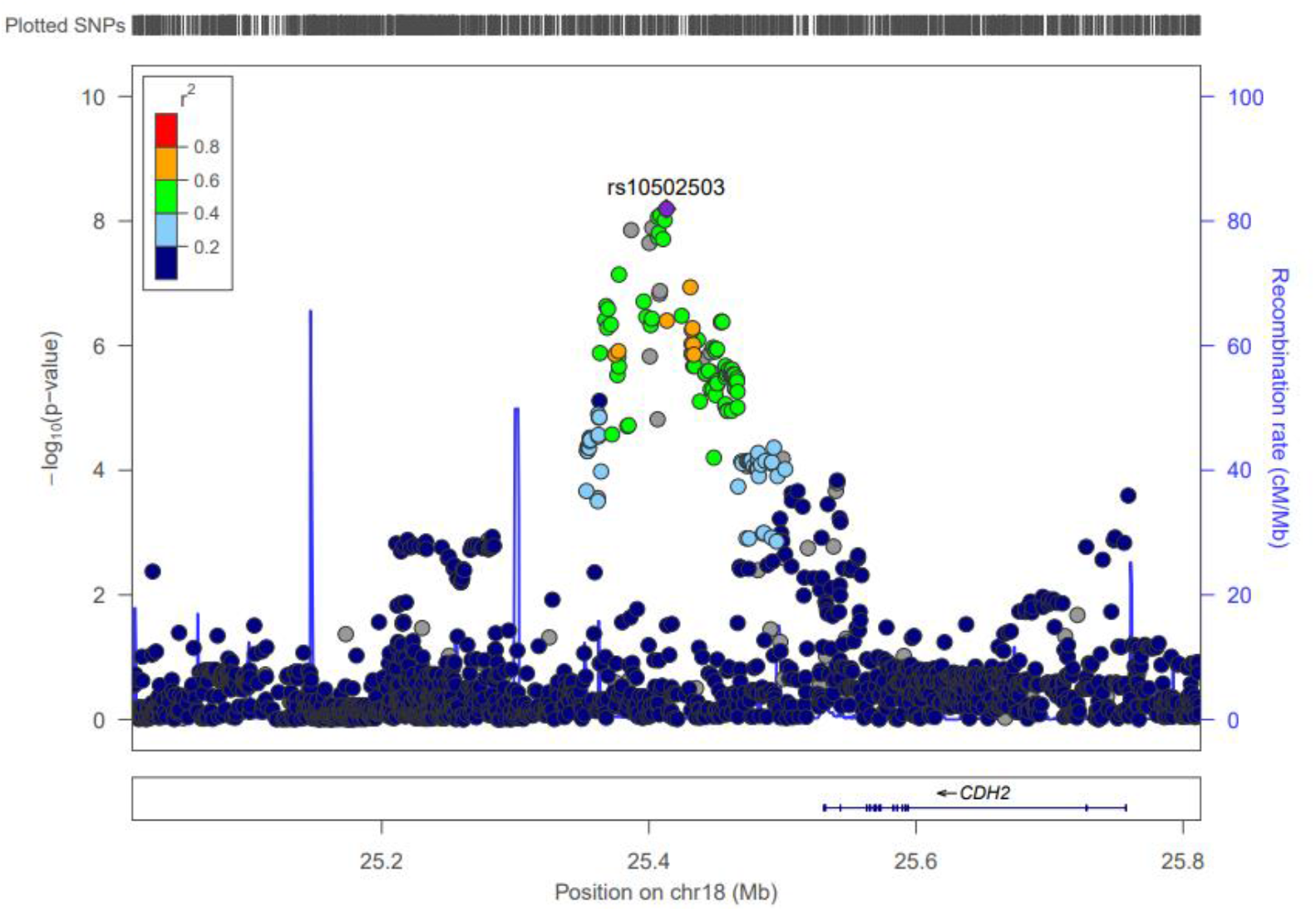
Regional plot for the chromosome 18q12.1 signal.

Functional annotation from FUMA mapped the association signal to *CDH2* on the basis of eQTL datasets including both multi-tissue samples (e.g., “eQTLgen_cis_eQTLs”) and cardiac and cerebellar tissues from GTEx data [28] (Supplementary Figure S5). Bayesian colocalization with default priors for eQTLs with at least 10 shared SNPs with PPD susceptibility indicated evidence of shared causal variants for cerebellum from GTEx v7 (PP H4 = 92.6%), cerebellum from GTEx v8 (PP H4 = 93.4%) cerebellar hemisphere from GTEx v8 (PP H4 = 95.3%) and macrophages (Alasoo_2018_ge_macrophage_naive, PP H4 = 96.6%) (extended colocalization results available on Supplementary Table S2). While the latter results should be interpreted cautiously due to the large differences in coverage between summary statistics included in colocalization analysis -follow-up validation is needed-, it is worth noting that the cerebellar eQTL finding seems consistent with the documented role of CDH2 in neural migration in cerebellum [29, 30]. *CHST9* was also included in FUMA’s report, although it is not discussed next as the mapping evidence was based on a few significant eQTL markers from one study, which is unlikely to constitute strong evidence of shared causal variation based on colocalization. Notably, data retrieved through FUMA indicated opposite direction of effects between the current PPD GWAS and expression levels from *CDH2* eQTLs (Supplementary Figure S6). Namely, alleles with a decreasing beta coefficient for PPD (protective alleles) would generally increase beta coefficient for *CDH2* expression.

Next, several GWAS and phenome-wide association study (PheWAS) catalogues were accessed to query the lead variant above (rs10502503) in relation to different traits based on peer-reviewed research articles and other available data sources: Open Targets Genetics (https://genetics.opentargets.org, [31, 32]), Atlas of GWAS Summary Statistics (https://atlas.ctglab.nl, [33]), GWAS Catalog (https://www.ebi.ac.uk/gwas, [34]) and Oxford Brain Imaging Genetics Server (https://open.win.ox.ac.uk/ukbiobank/big40/pheweb33k, [35]). Overall, no PheWAS association result for rs10502503 would survive a stringent Bonferroni adjustment for multiple testing. With a lenient significance threshold of nominal *p*<10^−5^, resting heart rate [36] was the only association retrieved out of all mentioned databases; notably, genetic signals for this marker in the current PPD analysis and the external cardiac GWAS seem to be driven by different causal (yet unknown) variants (Supplementary Figure S7). Other associations in the vicinity of rs10502503 retrieved by FUMA through the GWAS Catalog are included in Supplementary Table S2.

RVAS was conducted on up to 9,449 cases and 134,521 controls of European ancestry within the subset of UK Biobank participants with exome sequencing data. Association test results were available for 787,363 single variants (99,147 solved through conventional logistic regression and 688,216 using Firth’s approach) and 17,958 gene collapse genotypes (16,364 standard regression tests and 1,594 computed with Firth’s method). No result from this stage was statistically significant after multiple testing adjustments (Supplementary Figure S8).

## Discussion

In this work, PPD was phenotyped in women from the UK Biobank by integrating information from medical histories, hospital encounters, general practitioner records and self-reported questionnaires, to conduct a GWAS of common variants in subjects of European-ancestry. Genetic association results revealed one statistically significant locus at chromosome at 18q12.1, indexed by rs10502503. Follow-up functional mapping of the results provided suggestive evidence of *CDH2* as candidate causal gene.

The findings should be interpreted with close attention to the pipeline used for PPD case-control phenotyping and to demographic characteristics of UK Biobank, particularly regarding prevalence of PPD. With the current diagnostic definitions, we identified 14,810 cases and 217,199 controls; this gives a prevalence estimate of 6.38%, which is lower than other documented estimates even in cohorts without a previous history of depression (e.g., 10-15% or higher [37]) or in the general population from the United Kingdom (e.g., 21.5% [1]), likely reflecting a known “healthy volunteer” selection bias in the UK Biobank [38]. Notwithstanding this observation, medical risk factors observed in PPD cases were consistent with existing epidemiological literature (Table 1), and effect sizes from the genetic association results (odds ratio around 0.92) are similar to the ones typically reported for common variants in GWAS of common diseases and common variants.

To the knowledge of authors, the current analysis is the largest PPD GWAS conducted to date, and the outcomes include the first genome-wide significant locus reported for this disorder. As described in the introduction, related literature reports include a recent perinatal depression GWAS of 3,804 cases and 6,134 controls by Kiewa and colleagues [11], where no GWAS-significant hit was observed. It is worth highlighting that phenotyping from the latter study was done using the Edinburgh Postnatal Depression Scale, a dedicated psychometric scale to detect PPD with likely higher accuracy than the pipeline adopted for this study (using medical diagnoses and self-reported questionnaires). A strength of this study is the substantially larger numbers of cases and controls used for GWAS. However, we also recognize that the PPD definition in our study is limited by the ICD-derived diagnoses and self-reported information available, and does not rely on the more rigorous psychometric scale mentioned above. Notably, beyond these differences in clinical criteria, phenotypes considered here also diverge from the former GWAS in that here we consider mainly postnatal traits, whereas the former study’s authors considered the broad perinatal period (from beginning of pregnancy to six months post-delivery) [11].

Regarding how GWAS outcomes might be affected by the previous between-study phenotype differences -and by the potential non-specificity of the current PPD definition-, it has been noted that genetic association analyses conducted even with minimal phenotyping can identify susceptibility loci provided that sample sizes are large enough (e.g., self-reports of being a morning person can provide insights on the genetics of chronotype [39]), but research also suggests that minimal phenotyping of psychiatric traits can yield genetic signals of low specificity [40]. As with most genetic association findings, multiple stages of replication using independent samples of the same and other ancestries phenotyped through different instruments, and examination through follow-up experiments, are necessary steps to better understand the biological relevance of a given locus. Similarly related to phenotyping limitations, it is worth mentioning that, since the UK Biobank is a registry-based cohort and the diagnostic pipeline relied on ICD codes and self-reports, the extent of disease heterogeneity and the likelihood of misclassifications cannot be accurately estimated. Different datasets from PPD patients identified using psychometric tools for PPD and phenotyped directly at clinical settings might assist in confirming and expanding the present findings.

Post-GWAS analyses of the chromosome 18q12.1 signal suggested *CDH2* as candidate causal gene for PPD. Notwithstanding the limitations of existing GWAS fine-mapping and gene prioritization strategies [41], and despite not finding clear overlapping association signals (pleiotropy) for rs10502503 in different GWAS catalogues of common genetic variation, *CDH2* seems a plausible gene based on other sources of evidence. The CDH2 protein, also known as neural cadherin, is expressed in brain, heart and other tissues, plays a role in neural development, and *CDH2* genetic mutations can cause neurodevelopmental syndromic conditions characterized by intellectual disability and hypoplasia or agenesis of the corpus callosum [42, 43]. Similarly, in the neuropsychiatric domain, recent evidence indicates a link between protein maturation disruptions caused by missense *CDH2* mutations and attention-deficit hyperactivity disorder [44].

Interestingly, the locus at chromosome 18q12.1 identified in this study does not show strong links to other traits, as suggested by the lack of significant results in comprehensive catalogues of genotype-phenotype associations from common variants. Hypotheses derived from this observation are twofold. First, the apparent genetic specificity of the signal identified here might be related to findings on the genetic architecture of PPD in twins (*Introduction* section) [9], which indicated that a significant fraction of PPD heritability is not shared with the general depression phenotype. Hence, *CDH2* -or other pathways derived from the 18q12.1 signal-could point to genes and biological processes conferring susceptibility to PPD but not for other mental health disorders. Second, the association reported here indicates that the C allele of rs10502503 can protect against PPD and has a general lack of pleiotropy with other traits, which might be interpreted as a case of genetic modularity [45, 46]. The concept of modularity refers to a variational property that allows different characters to be expressed independently, an advantageous evolutionary strategy that permits optimization of each character by selection, without interference [45]. Genetic modularity can influence evolvability, understood as the capacity of an organism to produce phenotypic variation that is both adaptive and heritable [45, 47], and the finding for rs10502503 and PPD might be indicative of constrained genetic factors that evolved to boost maternal fitness.

Overall, this analysis identified a novel candidate locus associated with reduced risk for PPD and suggest the *CDH2* gene and its related biological pathways in disease etiology. Leveraging carefully curated medical records and self-reported mental health histories from large-scale public databases offers an unprecedented opportunity to research the genetic architecture of PPD to ultimately influence therapeutic interventions.

## Data Availability

Data from UK biobank cannot be shared openly and are subject to UK Biobank ethical approval. Further information about applying for UK Biobank data access can be obtained from the UK Biobank website (https://www.ukbiobank.ac.uk) or by emailing UK Biobank.

## Acknowledgements

This research has been conducted using the UK Biobank Resource under Application Number 26041. Under UK Biobank Application #26041, this work uses data provided by patients and collected by the NHS as part of their care and support (Copyright © (2022), NHS England. Re-used with the permission of the NHS England and UK Biobank. All rights reserved.). Under UK Biobank Application #26041, this work also uses data assets made available by National Safe Haven as part of the Data and Connectivity National Core Study, led by Health Data Research UK in partnership with the Office for National Statistics and funded by UK Research and Innovation (research which commenced between 1st October 2020 – 31st March 2021 grant ref MC_PC_20029; 1st April 2021 -30th September 2022 grant ref MC_PC_20058).

## Funding

Takeda Development Center Americas, Inc. provided funding. They had no role in study design, data analysis or preparation of the manuscript.

## Competing interests

ACP, DD and SS are employees of Takeda Development Center Americas, Inc. DD and SS own stock/stock options in Takeda. DD is shareholder of Merck, Sharp and Dohme. SS is shareholder of Johnson & Johnson.

## Notes

### Author Declarations

This research has been conducted using the UK Biobank Resource under Application Number 26041. Under UK Biobank Application #26041, this work uses data provided by patients and collected by the NHS as part of their care and support (Copyright (2022), NHS England. Re-used with the permission of the NHS England and UK Biobank. All rights reserved.). Under UK Biobank Application #26041, this work also uses data assets made available by National Safe Haven as part of the Data and Connectivity National Core Study, led by Health Data Research UK in partnership with the Office for National Statistics and funded by UK Research and Innovation (research which commenced between 1st October 2020 31st March 2021 grant ref MC_PC_20029; 1st April 2021 30th September 2022 grant ref MC_PC_20058).

