## Supplementary data for "Genome-wide Association Study of Postpartum Depression Identifies a Novel Susceptibility Locus at 18q12.1"

***Supplementary Material***

The following information refers to the manuscript ***Genome-wide Association Study of Postpartum Depression Identifies a Novel Susceptibility Locus at 18q12.1***.

| ***Item*** | ***Page*** |
|---|---|
| Supplementary Figure S1 | 2 |
| Supplementary Figure S2 | 3 |
| Supplementary Figure S3 | 4 |
| Supplementary Figure S4 | 5 |
| Supplementary Table S1 | 6 |
| Supplementary Figure S5 | 7 |
| Supplementary Table S2 | 8 |
| Supplementary Figure S6 | 9 |
| Supplementary Figure S7 | 10 |
| Supplementary Table S3 | 11 |
| Supplementary Figure S8 | 12 |

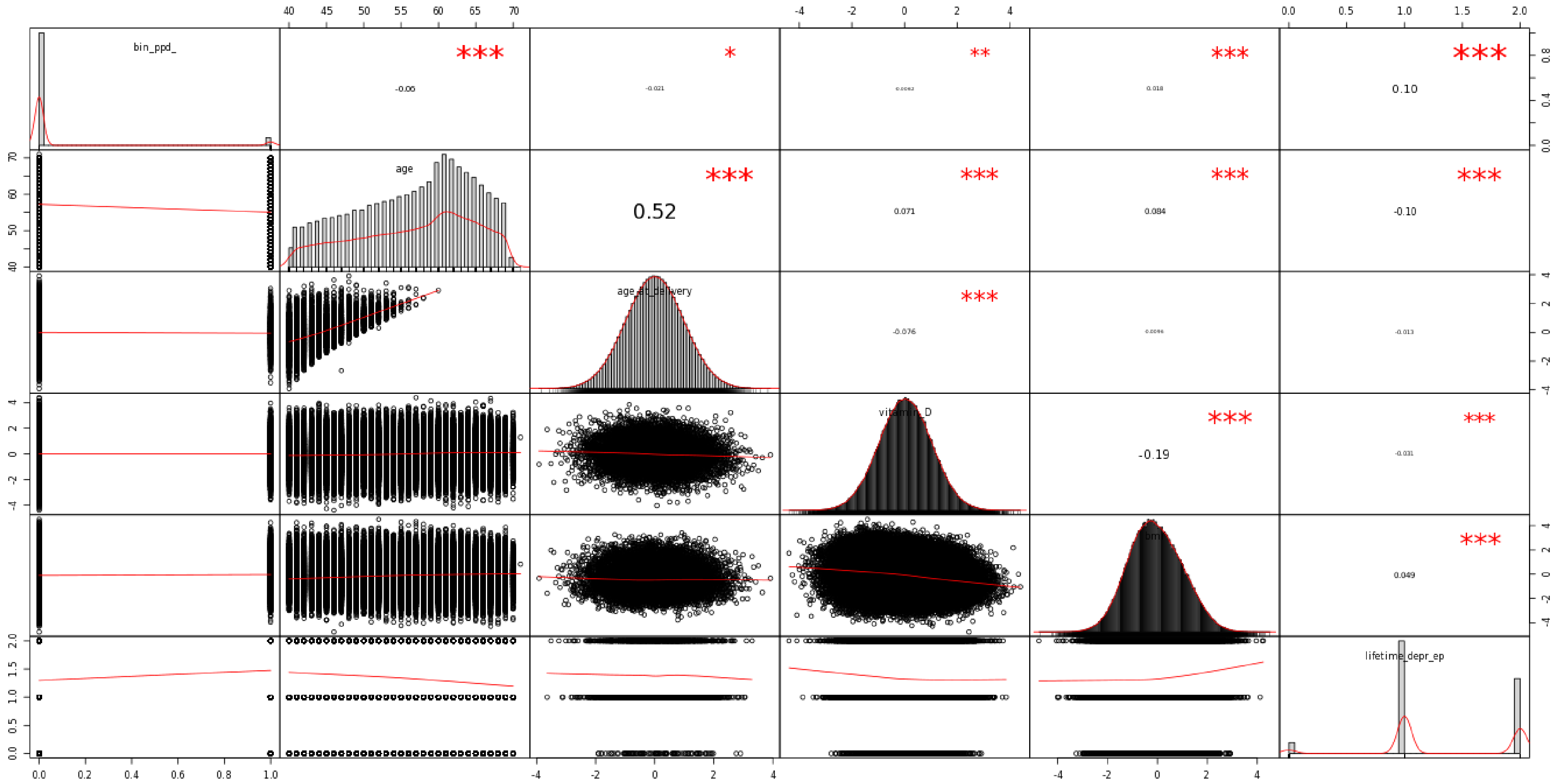

**Supplementary Figure S1.** Correlations between participants' diagnose, age, age at delivery, vitamin D levels, body mass index and number of self-reported depression episodes.

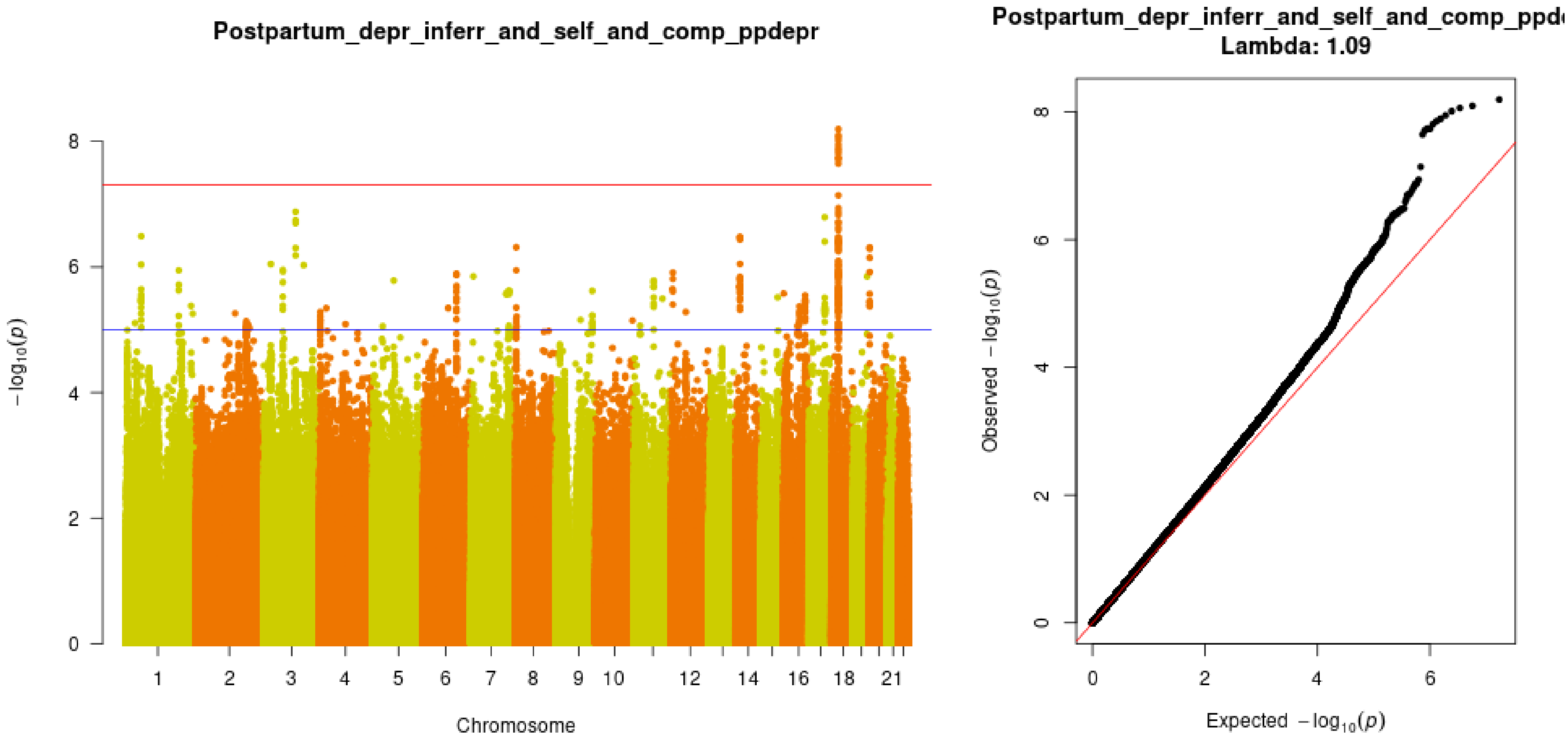

**Supplementary Figure S2.** Manhattan and QQ-plots from the PPD GWAS, including lambda genomic control estimate.

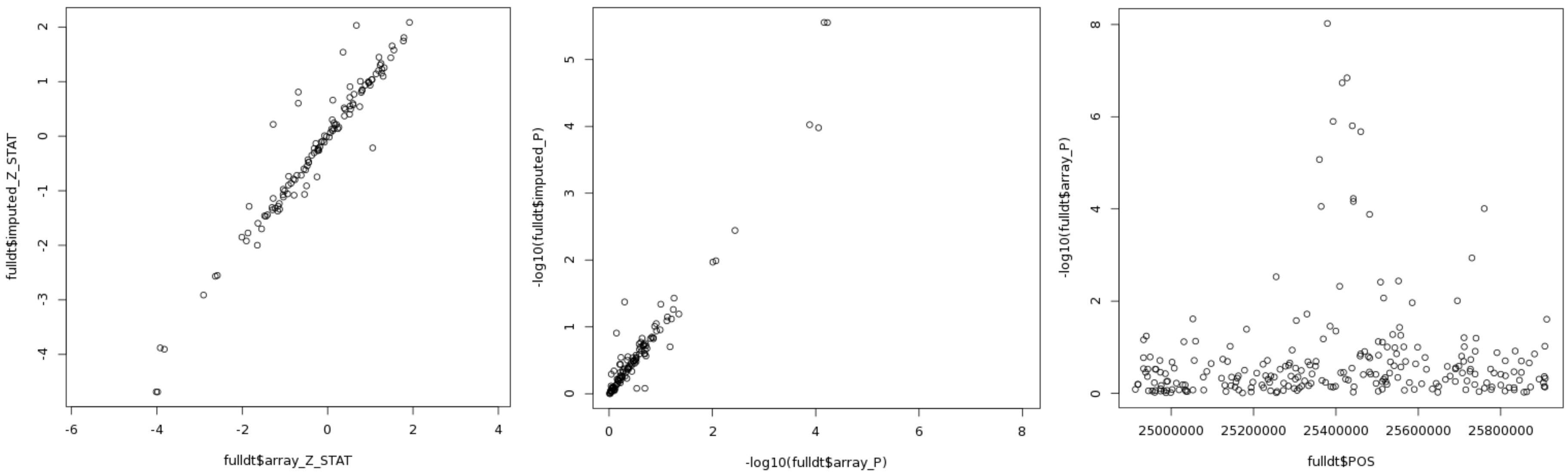

**Supplementary Figure S3.** Comparison between association results from imputed and directly genotyped markers, at the candidate chr18 locus.

Left: Z-statistic from array and imputed genotypes. Center: minus log-10 p-values from array and imputed genotypes. Right: regional association plot from directly genotyped data only.

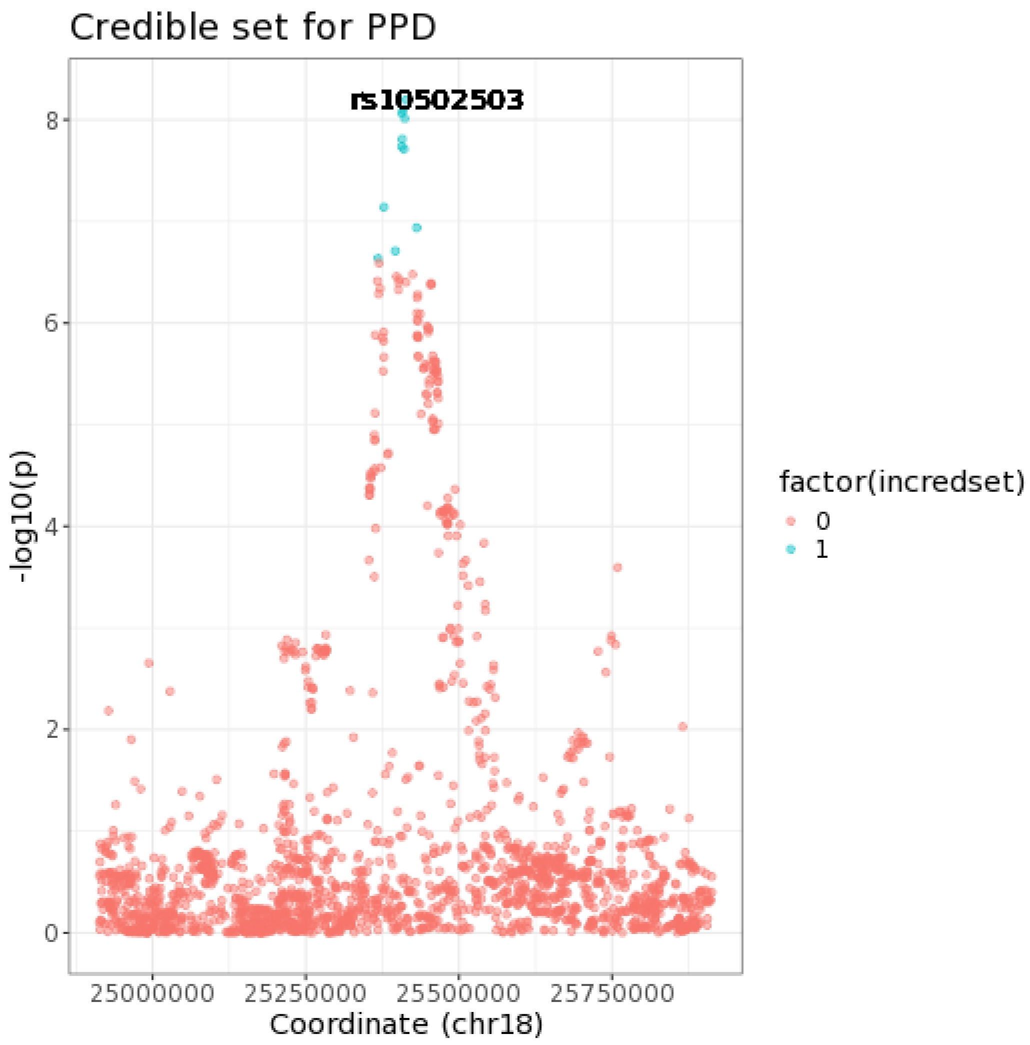

**Supplementary Figure S4.** Credible set of variants on PPD association locus near *CDH2*, as calculated using Bayesian fine-mapping.

| chr | bp | snp | PMID | FirstAuth | Study | Trait | ReportedGene | Strongest | Context | RiskAF | P | OrBeta | 95CI |
|---|---|---|---|---|---|---|---|---|---|---|---|---|---|
| 18 | 25406855 | rs9959491 | 29059683 | Michailidou K | Association analysis identifies 65 new breast cancer risk loci. | Breast cancer | NR | rs9959491-T | intergenic | 0.7115 | 5.00E-07 | 0.0345 | [0.021-0.048] unit increase |
| 18 | 25457298 | rs4355011 | 29534301 | Nishida N | Key HLA-DRB1-DQB1 haplotypes and role of the BTNL2 gene for response to a hepatitis B vaccine. | Response to hepatitis B vaccine | CDH2 | rs4355011-? | intergenic | | 7.00E-06 | 0.122 | unit increase |
| 18 | 25461745 | rs11083232 | 31484785 | Wright KM | A Prospective Analysis of Genetic Variants Associated with Human Lifespan. | Parental longevity (mother's age at death) | NR | rs11083232-G | intergenic | NR | 4.00E-06 | 0.01294 | [0.0074-0.0184] unit decrease |
| 18 | 25475798 | rs4258701 | 31511532 | Teumer A | Genome-wide association meta-analyses and fine-mapping elucidate pathways influencing albuminuria. | Urinary albumin-to-creatinine ratio in diabetes | CDH2 | rs4258701-T | intergenic | 0.3272 | 1.00E-08 | 0.0385 | [0.025-0.052] unit increase |

**Supplementary Table S1.** GWAS catalog associations for variants neighboring the lead PPD association (rs10502503). Data automatically reported by FUMA.

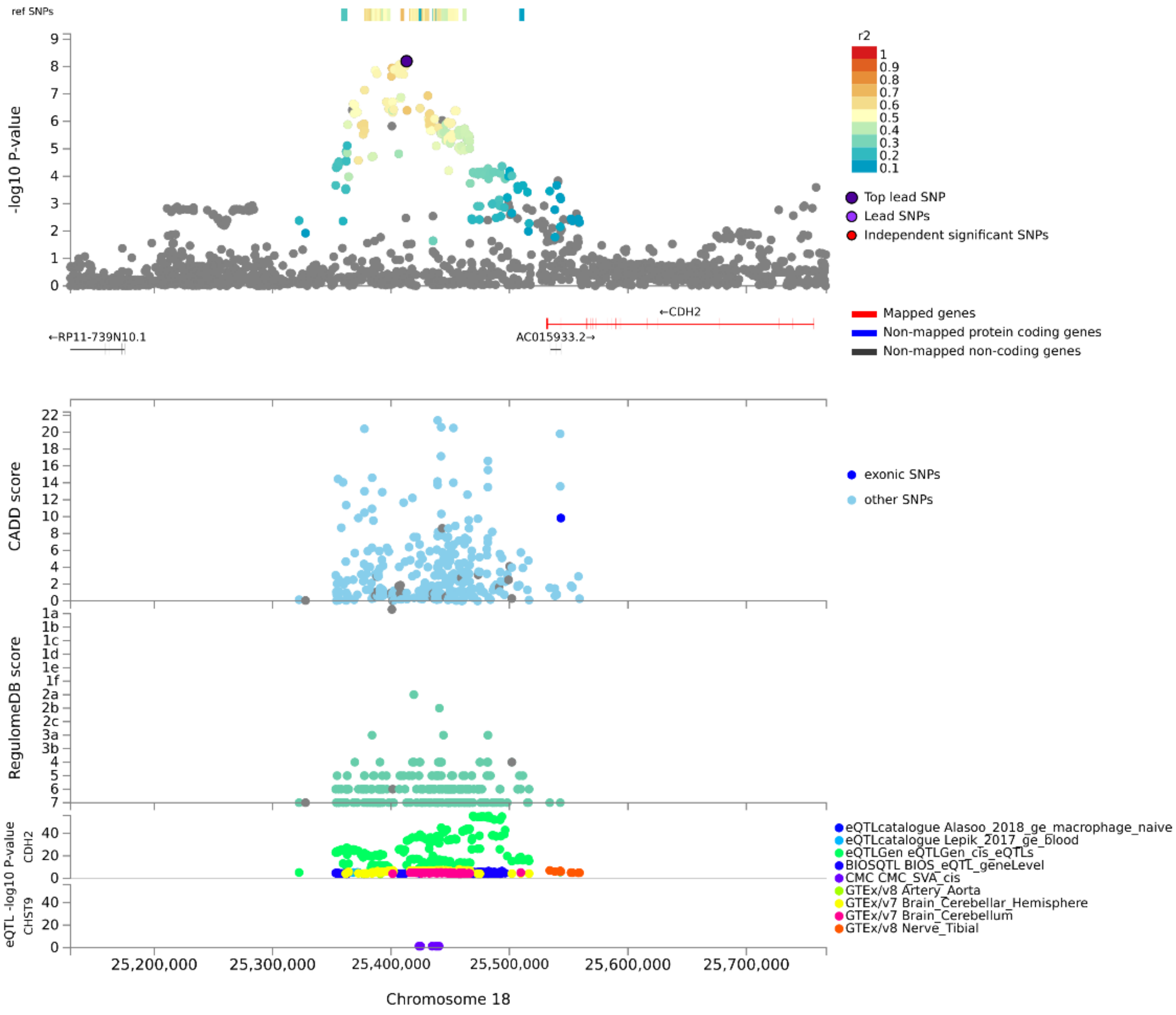

**Supplementary Figure S5.** Regional association plot retrieved from FUMA, displaying CADD and RegulomeDB scores, and eQTL data across multiple tissues.

| | nsnps | PP.H0.abf | PP.H1.abf | PP.H2.abf | PP.H3.abf | PP.H4.abf |
|---|---|---|---|---|---|---|
| **eQTLcatalogue_Alasoo_2018_ge_macrophage_naive_CDH2** | 25 | 3.40E-04 | 1.25E-02 | 0.000598 | 0.021005 | 0.965564 |
| **eQTLGen_eQTLGen_cis_eQTLs_CDH2** | 141 | 5.66E-53 | 3.90E-50 | 0.001416 | 0.974781 | 0.023803 |
| **BIOSQTL_BIOS_eQTL_geneLevel_CDH2** | 70 | 2.90E-05 | 1.20E-02 | 0.000901 | 0.372718 | 0.614348 |
| **GTEx.v8_Brain_Cerebellar_Hemisphere_CDH2** | 54 | 4.21E-05 | 1.10E-03 | 0.001715 | 0.043829 | 0.953315 |
| **GTEx.v8_Brain_Cerebellum_CDH2** | 37 | 1.91E-03 | 2.58E-02 | 0.002396 | 0.031549 | 0.938306 |
| **GTEx.v7_Brain_Cerebellum_CDH2** | 16 | 9.00E-03 | 4.84E-02 | 0.002802 | 0.014131 | 0.925715 |

**Supplementary Table S2.** Bayesian colocalization results for eQTLs retrieved from FUMA.

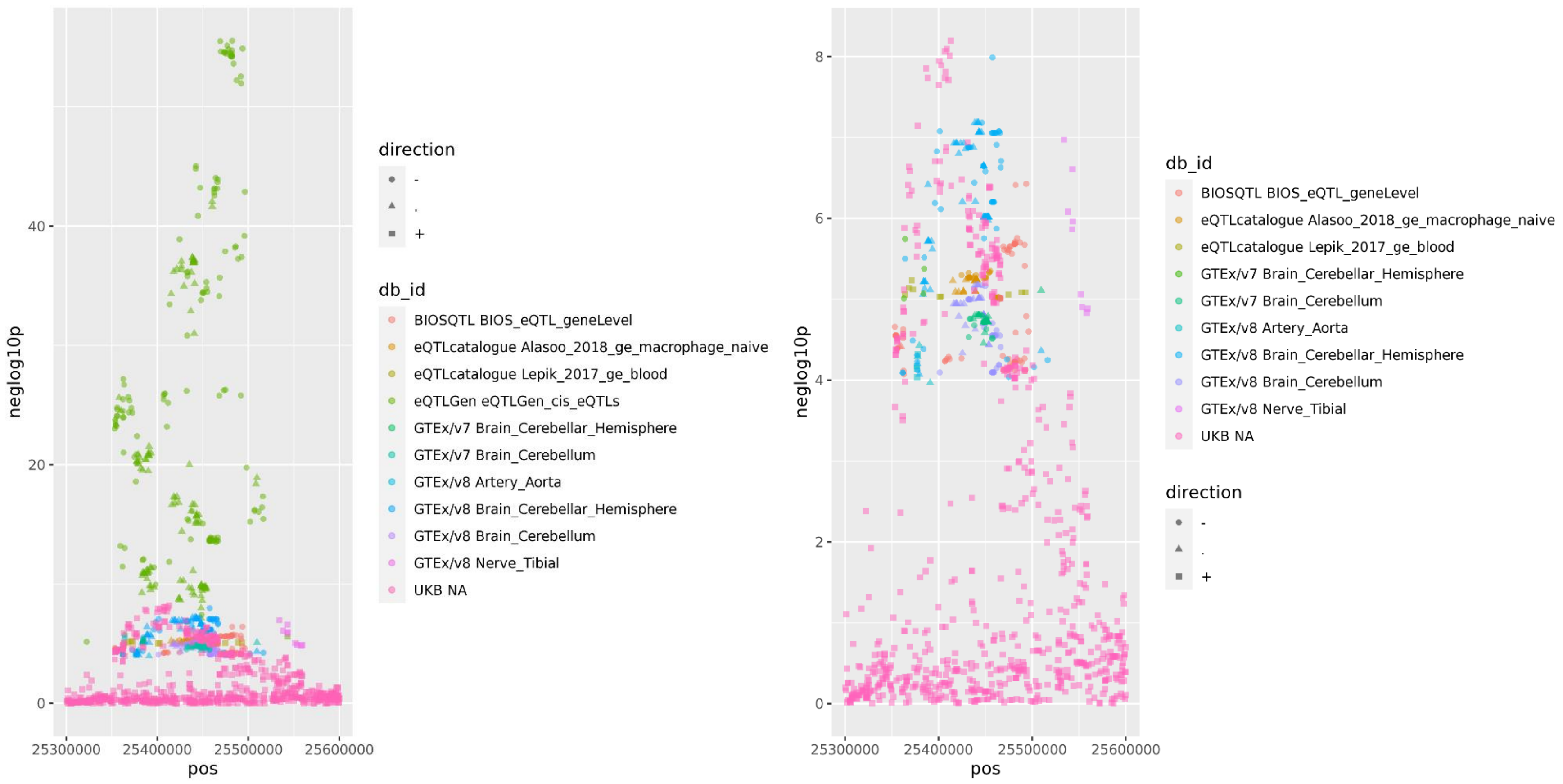

**Supplementary Figure S6.** Genomic region displaying an association with PPD, with overlaid eQTL information for *CDH2* from FUMA.

Notes: left, all retrieved eQTL datasets are shown; right, for clarity, “eQTLGen eQTLGen_cis_eQTLs” is omitted from the plot.

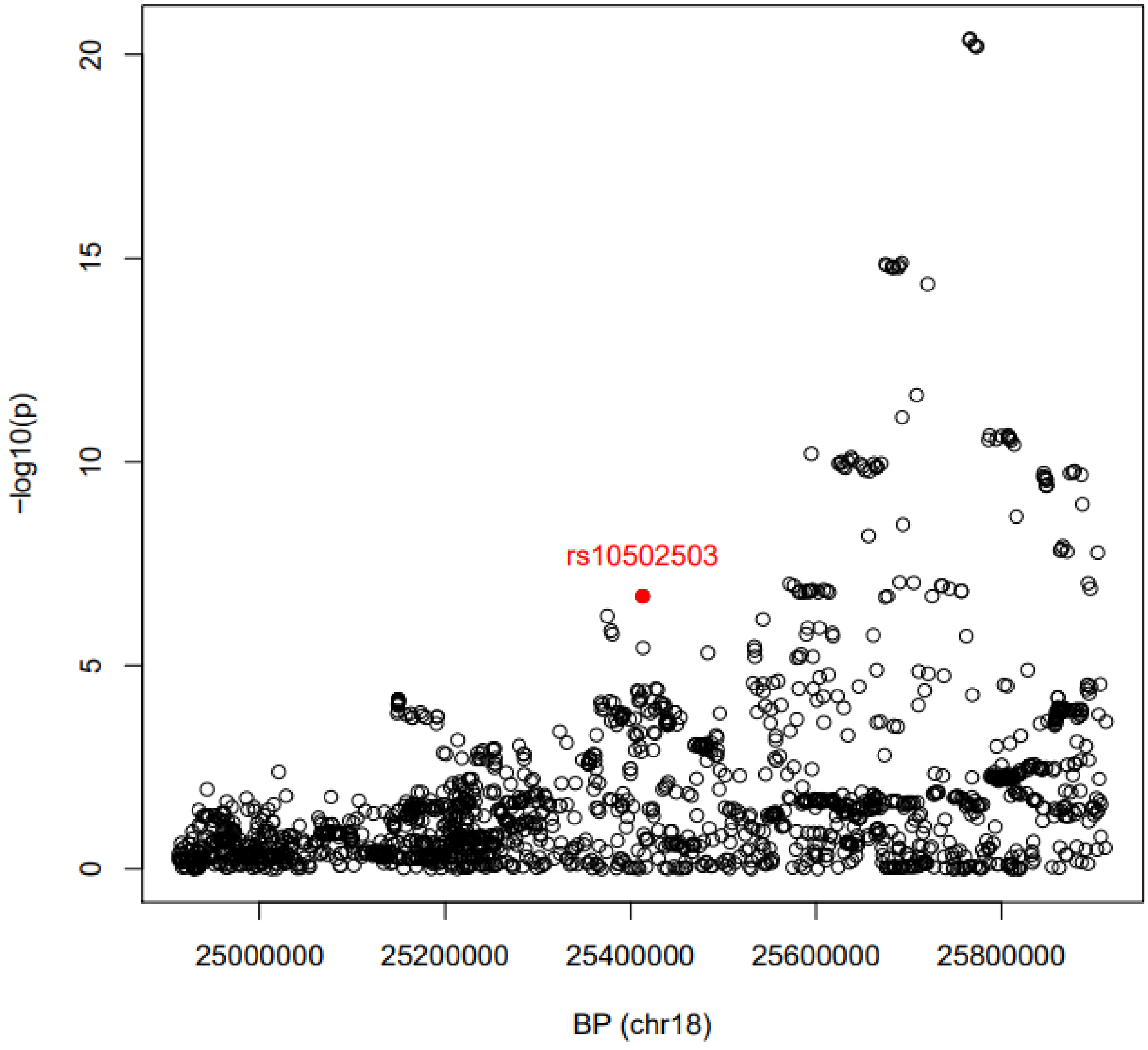

**Supplementary Figure S7.** Regional plot for the genetic association of resting heart rate around the lead PPD variant (rs10502503).

Note: summary statistics were retrieved from Zhu, Z., et al., Genetic overlap of chronic obstructive pulmonary disease and cardiovascular disease-related traits: a large-scale genome-wide cross-trait analysis. Respir Res, 2019. 20(1): p. 64.

| CHR | POS | ID | A1 | AX | A1_CASE_CT | A1_CTRL_CT | CASE_ALLELE_CT | CTRL_ALLELE_CT | A1_FREQ | OBS_CT | BETA | SE | Z_STAT | P |
|---|---|---|---|---|---|---|---|---|---|---|---|---|---|---|
| 18 | 25413104 | rs10502503 | C | T | 6645.16 | 100417 | 23564 | 334960 | 0.298618 | 179262 | -0.08732 | 0.015038 | -5.80629 | 6.39E-09 |
| 18 | 25408642 | rs11662671 | C | T | 6586.61 | 99462 | 23564 | 334960 | 0.295792 | 179262 | -0.08687 | 0.015063 | -5.76716 | 8.06E-09 |
| 18 | 25406866 | rs67108301 | T | C | 6588.06 | 99470.9 | 23564 | 334960 | 0.295821 | 179262 | -0.08668 | 0.01506 | -5.75529 | 8.65E-09 |
| 18 | 25411773 | rs2002401 | C | A | 6588.35 | 99449 | 23564 | 334960 | 0.295761 | 179262 | -0.08646 | 0.015074 | -5.73558 | 9.72E-09 |
| 18 | 25407513 | rs8092192 | C | G | 6593.06 | 99441.7 | 23564 | 334960 | 0.295754 | 179262 | -0.08515 | 0.015055 | -5.65621 | 1.55E-08 |
| 18 | 25406855 | rs9959491 | C | T | 6595.77 | 99449.6 | 23564 | 334960 | 0.295783 | 179262 | -0.08469 | 0.015051 | -5.62726 | 1.83E-08 |
| 18 | 25410603 | rs11876427 | G | A | 6576.17 | 99151.9 | 23564 | 334960 | 0.294898 | 179262 | -0.08467 | 0.015075 | -5.61684 | 1.94E-08 |
| 18 | 25377582 | rs16944182 | C | T | 5006.22 | 76165.5 | 23564 | 334960 | 0.226405 | 179262 | -0.08899 | 0.016524 | -5.38528 | 7.23E-08 |
| 18 | 25431060 | rs8093989 | T | C | 5587.55 | 84525.2 | 23564 | 334960 | 0.251344 | 179262 | -0.0843 | 0.015905 | -5.30018 | 1.16E-07 |
| 18 | 25396110 | rs7243670 | T | C | 9344.54 | 138484 | 23564 | 334960 | 0.412324 | 179262 | -0.07176 | 0.013791 | -5.20351 | 1.96E-07 |
| 18 | 25368076 | rs7235990 | C | A | 5415.28 | 81817.7 | 23564 | 334960 | 0.243311 | 179262 | -0.08335 | 0.016115 | -5.17189 | 2.32E-07 |

**Supplementary Table S3.** Summary statistics for variants in the credible set of locus near *CDH2*.

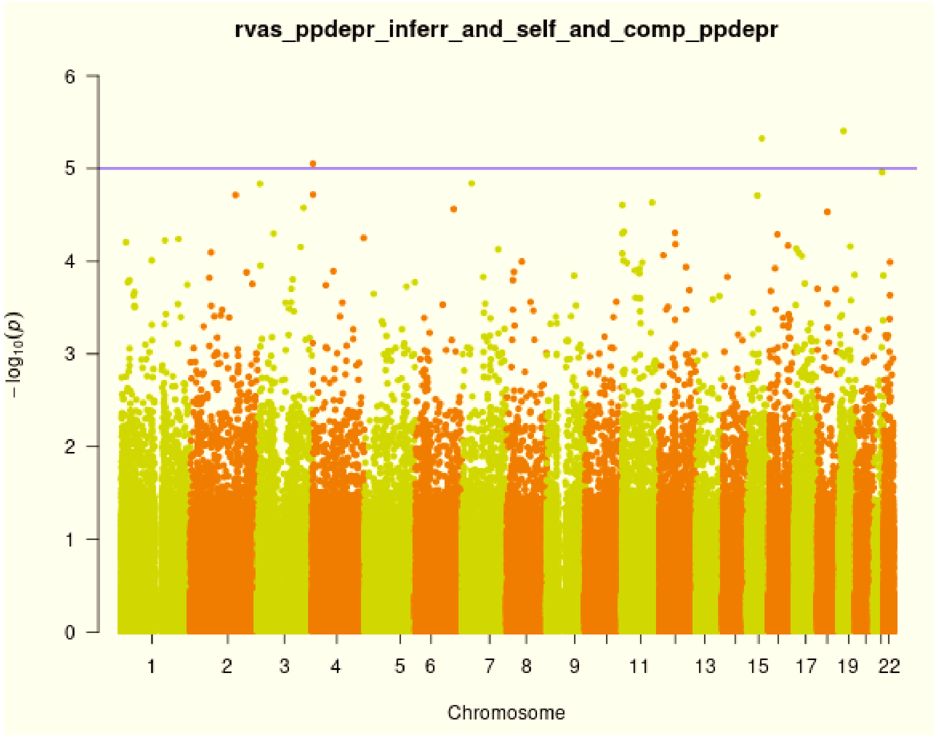

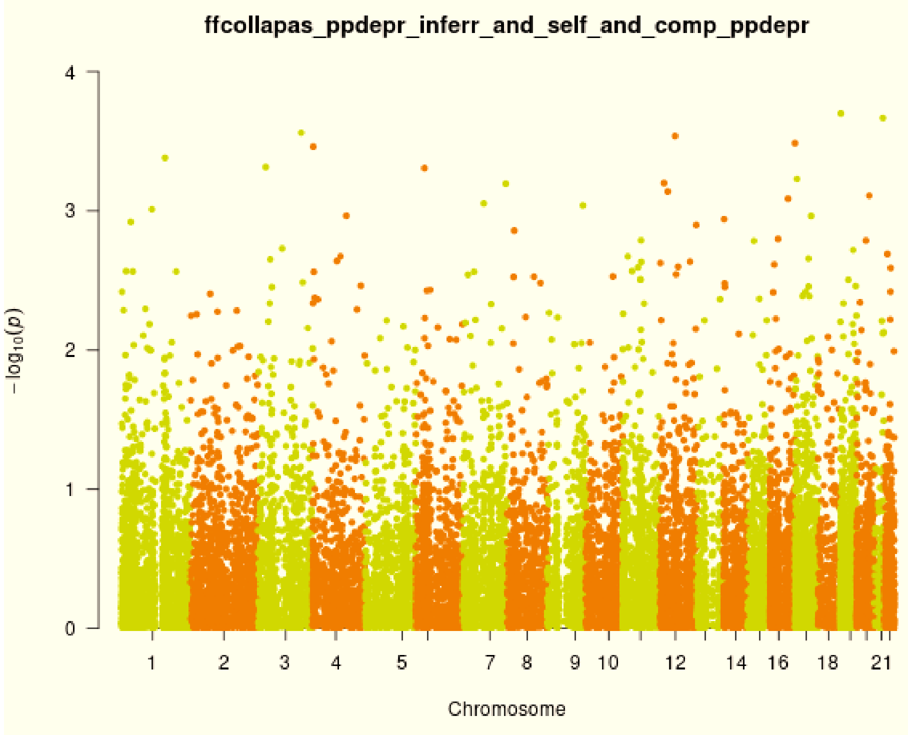

**Supplementary Figure S8.** Results of RVAS for single variants (left) and gene collapse genotypes (right).
